# Quantifying Exercise Intensity to Predict Changes in Walking Capacity in People with Chronic Stroke

**DOI:** 10.1101/2025.03.04.25323345

**Authors:** Kiersten M. McCartney, Pierce Boyne, Ryan Pohlig, Susanne M. Morton, Darcy Reisman

## Abstract

**Objective:** To examine if exercise intensity, quantified as heart rate or training speed, predicts walking outcomes in people with chronic stroke.

**Design:** This is a secondary analysis from a larger randomized clinical trial (“*PROWALKS*”; NIH1R01HD086362).

**Setting:** Four, outpatient rehabilitation clinics.

**Participants:** Participants with chronic stroke with a walking speed of 0.3-1.0m/s and step-activity of <8000 steps-per-day. This analysis included participants (*n* = 169; age: 63.1 ± 12.5, 46% female) with complete pre- and post-intervention data.

**Interventions:** Participants were randomized into (1) fast-walking training or (2) fast-walking training and step-activity monitoring group. Of importance, participants received up to 36-sessions of 30-minute high-intensity treadmill walking training across 12-weeks.

**Main Outcome Measure(s):** The primary outcomes were a pre-to-post intervention change in six-minute walk test distance and fastest walking speed. Exercise intensity was quantified as either a percentage of heart rate reserve or self-selected walking speed.

**Results:** Four, separate multiple linear regressions with robust errors analyzed the relationship of exercise intensity (*predictor;* % heart rate or training speed) on pre-to-post intervention changes in walking capacity outcomes (*outcome;* Six-Minute Walk Test, Fastest Walking Speed) after accounting for covariates.

Training speed was a significant predictor of both a change in Six-Minute Walk Test distance (*b* = 0.364 (95% CI [0.108 - 0.621]), *p* = 0.006) and Fastest Walking Speed (*b* = 0.001 (95% CI [0.001-0.002]), *p* = 0.003). Heart rate was not a significant predictor of either outcome (both *p* > 0.096).

**Conclusions:** Training speed significantly predicts changes in walking capacity outcomes in people with chronic stroke following. This suggests rehabilitation clinicians may use training speed as the metric of exercise intensity when prescribing walking interventions to people with chronic stroke.

## INTRODUCTION

More than 80% of people with chronic stroke experience a severe reduction in their ability to walk.^1^ This includes notable decreases in their walking capacity, collectively represented by walking speed and walking endurance.^1–4^ Moderate-to-high intensity walking interventions are currently the most effective intervention to improve walking capacity in individuals with chronic stroke.^5,6^ However, following these interventions, there remains significant variability in walking capacity outcomes across participants.^6^

The aggregation of aerobic and/or walking intervention studies in people with chronic stroke suggests that higher levels of intensity may facilitate greater changes in cardiorespiratory fitness, walking speed, and walking endurance.^7^ Exercise intensity is most frequently quantified as a percentage of heart rate maximum or heart rate reserve (HRR) in moderate-to-high walking interventions for people with chronic stroke. Heart rate is used as both the intensity metric which guides the exercise intervention design (e.g., “*training heart rates were established at 70-80% HRR”*)^8^ and the “real-time” response when a participant is exercising (e.g., “*Treadmill incline was adjusted or weight vests added as needed to keep heart rate in the target heart rate range during all walking*”).^9^ While heart rate is a valid and frequently used measure of cardiovascular intensity in response to exercise, it is influenced by other factors, agnostic to exercise, including stress, anxiety, sleep quality, and dehydration.^10,11^

The previous studies which have suggested higher intensity exercise predicts greater changes in walking outcomes in people with chronic stroke have had several challenges. These include small sample sizes, not accounting for important co-variates, and/or only demonstrating a correlation between intensity and outcome.^8,12–21^ Secondly, intensity has almost uniformly been quantified based on heart rate. This has limited our understanding to only one metric of exercise intensity.

While many individuals with chronic stroke have low cardiorespiratory fitness, they often also have significant neuromuscular deficits.^22,23^ These neuromuscular deficits may limit the cardiovascular intensity people with stroke can reach in a walking intervention. Therefore, it has been suggested that walking speed may provide a more specific measure of exercise intensity in this population.^24^ While it may be intuitive to assume that faster walking speeds are associated with or equated to higher heart rates, it is unknown if these metrics of exercise intensity are equally predictive of walking outcomes in people with chronic stroke. Probing which parameter of exercise intensity is most strongly predictive of walking outcomes remains largely unexplored.^24,25^ Understanding the strength of each exercise intensity metric is imperative to provide the optimal training parameters for clinicians to use when implementing walking exercise interventions in people with chronic stroke.

The purpose of this study was to examine how exercise intensity, quantified as either training heart rate or training speed, would impact changes in walking capacity outcomes in people with chronic stroke after accounting for several covariates. We hypothesized after accounting for important covariates, that exercise intensity quantified as either heart rate or training speed, would be a predictor of changes in walking endurance and fastest walking speed following a walking exercise intervention in people with chronic stroke.

## METHODS

### Participants

This is a secondary analysis from a larger randomized controlled trial “*Promoting Recovery Optimization of Walking Activity in Stroke”* (PROWALKS”; NIH 1R01HD086362). The full protocol and primary results have been published.^9,26^ All study procedures were approved by the University of Delaware Institutional Review Board and participants signed an informed consent prior to enrollment. Participants were 21-85 years old, had their most recent stroke at least 6 months prior to enrollment, had self-selected walking speeds of 0.3-1.0m/s, and could ambulate without the assistance of another person. Of the total 250 participants randomized to PROWALKS, only the 169 participants randomized into the (1) fast-walking training (FAST, *n* = 89) or (2) fast-walking training and step activity monitoring combined intervention (FAST + SAM, *n* = 80) were used for this analysis (Figure 1).^27^ The 81 participants randomized to the step activity monitoring intervention were excluded as they did not undergo any treadmill training.

**Figure 1.**
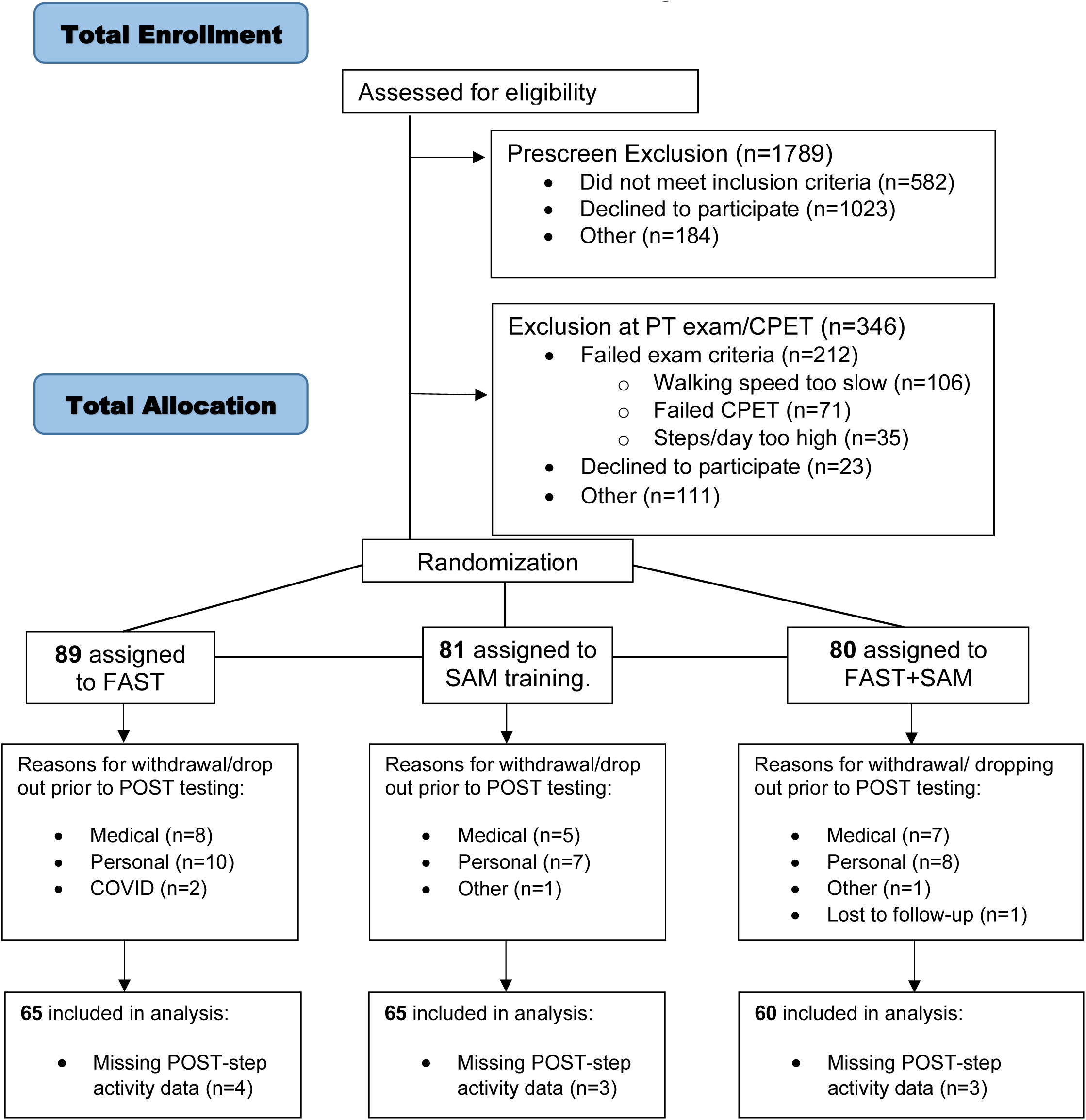
TOTAL CONSORT Flow Diagram.

### Clinical Evaluation

Prior to randomization, all participants underwent a baseline clinical evaluation which included collecting demographic and medical information. Demographic information included age and biological sex, and medical information included time since the most recent stroke (TSS) and the Charlson Comorbidity Index (CCI). The CCI is a 16-item questionnaire comorbidity scoring system which weights factors based on disease severity and quantifies comorbidity burden.^28,29^ Participants also completed the Activities Balance Confidence Scale (ABC), a reliable and valid 16-item self-report questionnaire that measures balance self-efficacy.^30,31^

Participants also completed measures of walking capacity. Walking capacity is defined as what a person *can do* usually as assessed by standardized tests conducted in structured environments such as a clinic or laboratory.^32^ The six-minute walk test (6MWT) and 10-meter walk test (10mWT) are valid measures and are recommended to quantify walking capacity in the chronic stroke population.^33,34^ The 6MWT may represent an individual’s walking endurance and be influenced by their cardiovascular fitness and/or neuromotor function.^33,35–37^ The 10mWT can capture individuals self-selected (SSWS) and fastest walking speeds (FWS) over a short distance.^33,38^ Participants were instructed to “*walk at your normal pace*” (SSWS) or “*walk at your fastest possible pace*” (FWS) and completed three trials at each walking speed, with the average of each trial recorded.^26^ Walking capacity measures were completed at baseline and after completing the intervention. The previously published protocol outlines all measures collected in the parent trial.^26^

### Exercise Intervention

Participants randomized to the FAST or FAST+SAM intervention received up to 36, 30-minute sessions of treadmill training over 12 weeks.^9,26^ The intervention protocol was designed to promote <u>continuous walking</u> at high cardiovascular intensities. The prescribed exercise intensity for all treadmill sessions was 70-80% of an individual’s heart rate reserve (HRR).^26^

Training physical therapists were free to manipulate treadmill speed, incline, and/or use ankle weights or a weighted vest to achieve the target heart rate zone. All participants wore a safety harness which did not provide any body weight support. The exercise session was terminated if a participant’s response violated the guidelines set forth by the American College of Sports Medicine (ACSM) for individuals in phase III or IV cardiac rehabilitation.^39^ Heart rate was monitored continuously (Polar H10 chest-straps), with heart rate and treadmill speed recorded each minute.

### Exercise Intensity

For this analysis exercise intensity was quantified in two ways: (1) the average percent HRR and (2) the average training speed attained across the entire intervention. Using the maximum heart rate (MHR) attained on the cardiopulmonary exercise stress test prior to randomization and the individual’s daily resting heart rate (RHR), percent HRR was quantified using the Karvonen method.^39^ Participants on beta-blocker medications were instructed to take their medications prior to the exercise test (which determined heart rate maximum) and for each training session. For each minute of heart rate (HR) data, percent HRR was calculated as:

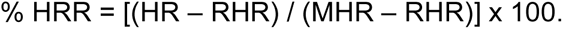

For each minute of treadmill speed data, training speed was calculated as a percentage of an individual’s baseline SSWS as follows:

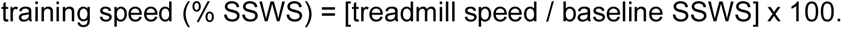

Prior to training speed calculations treadmill speeds were converted from miles per hour to meters per second.

For both exercise intensity metrics, values were averaged across every minute of training data from the intervention.

### Statistical Analyses

Four (4), separate multiple linear regressions with robust errors were run to analyze the relationship of exercise intensity (*predictor;* % HRR or training speed) on a baseline to post-intervention change in walking capacity outcomes (*outcome;* 6MWT, FWS) after accounting for covariates. Covariates included age, sex, TSS, CCI, ABC, baseline 6MWT, and total exercise volume. These covariates were chosen based on prior literature which has demonstrated their impact on rehabilitation outcomes or relationship to walking capacity in people with chronic stroke.^13,40–42^ If these covariates were not controlled for, it could signal a false relationship between exercise intensity and change in walking capacity outcome that is accounted for by other variables. Exercise volume was quantified as the total number of minutes walked in the intervention (e.g., if a participant attended 24 sessions and walked 26 minutes in each session, their exercise volume was 624 minutes). This was included to account for differences between participants in the number or frequency of training sessions and/or number of minutes walked within a session. Statistical analyses were conducted using Statistical Package for the Social Sciences (SPSS) (Version 29.0; IBM, Armonk, New York).

## RESULTS

One-hundred twenty-nine participants randomized to the FAST (*n* = 68) or FAST+SAM (*n* = 61) intervention completed both baseline and post-intervention walking capacity measures. Participants were (mean±SD) 63±13 years old, 46% female, were 45±56 months (∼3.75 years) post-stroke (Table 1), and had on average a 44m increase in 6MWT distance and a 0.13 and 0.17 m/s increase in SSWS and FWS, respectively (Table 2). Across the intervention, these participants attended 29 training sessions and walked for an average of 28 minutes per session at a heart rate intensity of 63.5% HRR (Table 3). There were no differences in baseline characteristics or training fidelity metrics between participants in the FAST or FAST+SAM interventions, so intervention group was not included as a covariate in the regression analyses. For clarity results are organized by exercise intensity predictor.

**Table 1:** Participant Characteristics.

| Characteristic | Participants (*n* = 129) |
| --- | --- |
| *Continuous* |  |
| Age (years) | 63.5 ± 12.5 |
| Time Since Initial Stroke (months) | 46.4 ± 57.8 |
| Body mass index (kg/m<sup>2</sup>) | 29.3 ± 6.3 |
| Self-selected walking speed (m/s) | 0.7 ± 0.2 |
| Fast walking speed (m/s) | 1.0 ± 0.3 |
| 6-Minute Walk Test (m) | 291.9 ± 117.2 |
| Average Baseline Steps/Day | 3,729.6 ± 1,862.6 |
| Activities Balance Confidence Scale | 74.0 ± 17.7 |
| Patient Health Questionnaire-9 | 3.7 ± 3.8 |
| Charlson Comorbidity Index | 3.3 ± 1.9 |
| *Categorical* |  |
| Sex | Male: *n* = 69 (53.5%)<br>Female: *n* = 60 (46.5%) |
| Side of Hemiparesis | Left: *n* = 63 (48.8%)<br>Right: *n* = 60 (46.5%)<br>Bilateral: *n* = 6 (4.7%) |
| Assistive Device Use (yes/no) | Yes: *n* = 60 (46.5%)<br>No: *n* = 69 (53.5%) |
| Orthotic Device Use (yes/no) | Yes: *n* = 34 (26.4%)<br>No: *n* = 95 (73.6%) |
| Ethnicity | Hispanic or Latino: *n* = 3 (4.7%)<br>Not Hispanic or Latino: *n* = 123 (95.3%) |
| Race | Native American/American Indian/Alaska Native: *n* = 5 (3.9%)<br>Asian: *n* = 10 (7.8%)<br>Native Hawaiian or Pacific Islander: *n* = 0 (0%)<br>African American/Black: *n* = 33 (25.6%)<br>Caucasian/White: *n* = 86 (66.7%)<br>Prefer not to Respond: *n* = 1 (0.8%)<br>More than 1 Race: Yes (*n* = 8, 6.2%) |
| Beta-Blocker Medication | Yes: *n* = 49 (38.9%)<br>No: *n* = 77 (61.1%) |

*Continuous data presented as mean ± SD; Categorical data presented as n (% sample)*

**Table 2:** Changes in Walking Capacity Outcomes.

| **Outcome** |  |
| --- | --- |
| **Six-Minute Walk Test (m)** |  |
| *Post* | 335.1 ± 120.1<br>(74.7 – 660.2) |
| *Change from Baseline* | 44.0 ± 44.5<br>(-66.2 – 198.2) |
| **Fastest Walking Speed (m/s)** |  |
| *Post* | 1.17 ± 0.41<br>(0.27 – 2.1) |
| *Change from Baseline* | 0.17 ± 0.17<br>(-0.13 - 0.73) |
| **Self-Selected Walking Speed (m/s)** |  |
| *Post* | 0.84 ± 0.27<br>(0.26 – 1.48) |
| *Change from Baseline* | 0.13 ± 0.13<br>(-0.14 - 0.52) |

*Data presented as mean ± SD (range)*

**Table 3:** Exercise Dose Parameters.

| **Exercise Dose Parameter** |  |
| --- | --- |
| **Number of Sessions** | 29.0 ± 4.1<br>(19-36) |
| **Volume of Walking (minutes)** | 823.0 ± 130.2<br>(427 – 1050) |
| **Average Heart Rate Reserve (%)** | 63.6 ± 12.3<br>(19.6 – 123.3) |
| **Average Training Speed (% of SSWS)** | 107.0 ± 32.0<br>(36.3 – 260.6) |

*Data presented as mean ± SD (range).*
*SSWS = self-selected walking speed. If a participant walked for all 30 minutes at each of a* *maximum of 36 sessions, the highest possible exercise volume was 1080 minutes.*

### Exercise Intensity: Heart Rate Reserve

#### Outcome – Change in 6MWT

The model was not significant (R^2^ = 0.077, F(8,118) = 1.237, *p* = 0.284) (Supplemental Table 1).

#### Outcome – Change in FWS

The model was significant (R^2^ = 0.132, F(8,119) = 2.261, *p* = 0.028). Average percent HRR was not a significant predictor (*b* = -0.001 (95% CI: -0.003 - 0.002), *p* = 0.546) of change in FWS after accounting for all covariates (Table 4a).

**Table 4a:** Heart Rate Reserve as a Predictor of Change in FWS.

| Parameter | *B* | Std. Error | *t* | Sig. | 95% CI |
| --- | --- | --- | --- | --- | --- |
| Intercept | 0.278 | 0.206 | 1.350 | 0.180 | -0.130 – 0.685 |
| Sex | 0.005 | 0.032 | 0.147 | 0.884 | -0.059 – 0.069 |
| Age | -0.001 | 0.001 | -0.922 | 0.358 | -0.003 – 0.001 |
| TSS | 0.000 | 0.000 | -1.417 | 0.159 | -0.001 – 0.000 |
| Baseline 6MWT | 0.000 | 0.000 | 2.539 | 0.012 | 0.000 – 0.001 |
| ABC | 0.001 | 0.001 | 1.182 | 0.239 | -0.001 – 0.003 |
| CCI | -0.007 | 0.007 | -1.001 | 0.319 | -0.020 – 0.007 |
| Exercise Volume | 0.000 | 0.000 | -1.180 | 0.240 | 0.000 – 0.000 |
| Heart Rate Reserve | -0.001 | 0.001 | -0.606 | 0.546 | -0.003 – 0.002 |

### Exercise Intensity: Training Speed

#### Outcome – Change in 6MWT

The model was significant (R^2^ = 0.125, F(8,118) = 2.115, *p* = 0.040). Average training speed was a significant predictor (*b* = 0.362 (95% CI [0.107 - 0.617]), *p* = 0.006) of change in 6MWT after accounting for all covariates (Table 4b).

**Table 4b:** Training Speed as a Predictor of Change in 6MWT.

| Parameter | *B* | Std. Error | *t* | Sig. | 95% CI |
| --- | --- | --- | --- | --- | --- |
| Intercept | 11.951 | 38.556 | 0.310 | 0.757 | -64.400 – 88.302 |
| Sex | 3.487 | 8.509 | 0.410 | 0.683 | -13.363 – 20.337 |
| Age | -0.225 | 0.294 | -0.766 | 0.445 | -0.806 – 0.356 |
| TSS | -0.046 | 0.050 | -0.917 | 0.361 | -0.146 – 0.053 |
| Baseline 6MWT | -0.072 | 0.040 | -1.810 | 0.073 | -0.150 – 0.007 |
| ABC | 0.252 | 0.312 | 0.805 | 0.422 | -0.367 – 0.870 |
| CCI | -0.969 | 2.537 | -0.382 | 0.703 | -5.994 – 4.055 |
| Exercise Volume | 0.013 | 0.029 | 0.437 | 0.663 | -0.045 – 0.070 |
| Training Speed | 0.362 | 0.129 | 2.815 | 0.006 | 0.107 – 0.617 |

#### Outcome – Change in FWS

The model was significant (R^2^ = 0.192; F(8, 119) = 3.530, *p* = 0.001). Average training speed was a significant predictor (*b* = 0.001 (95% CI [0.001-0.002]), *p* = .003) of change in FWS after accounting for all covariates (Table 4c).

**Table 4c:**
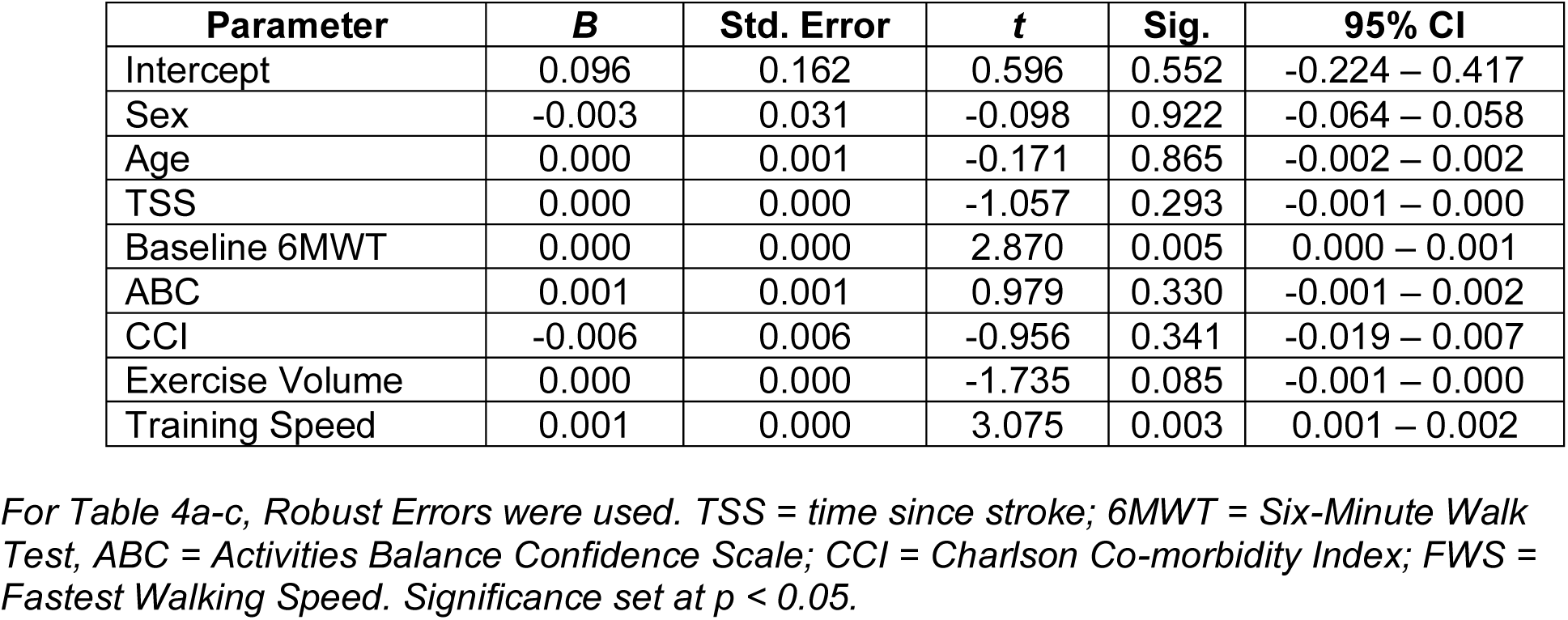
Training Speed as a Predictor of Change in FWS.

## DISCUSSION

This study sought to understand if training intensity (quantified as average percent HRR or training speed) is predictive of changes in walking capacity outcomes after accounting for multiple known co-variates following a moderate-to-high intensity intervention in people with chronic stroke. After accounting for covariates, training speed was a significant predictor of changes in both walking endurance (6MWT) and FWS. Conversely, training heart rate was not a significant predictor of change in either walking capacity outcome. With the largest known sample of individuals with chronic stroke to complete a fast-walking training intervention (*n* = 129), this analysis had the statistical power to account for covariates which may influence walking capacity outcomes. These results provide evidence of training speed as a predictor of changes in walking capacity outcomes *after* accounting for additional influential factors, such as age, baseline walking endurance, or balance self-efficacy.^12,15,43^ It was surprising that average training heart rate was not a predictor of walking capacity outcomes after accounting for known co-variates.

To put into a clinical perspective the impact of training at faster speeds, if participants walked on average at their SSWS, there was a 41.3m increase in 6MWT and .157m/s increase in FWS following the intervention. If participants walked on average at their FWS (∼133% of SSWS), there was a 58.0m increase in 6MWT and a 0.203m/s increase in FWS. On average, training at FWS led to greater changes in 6MWT distance (+16.7m) and FWS (+0.046m/s) compared to training at SSWS. This suggests challenging people with chronic stroke to walk at the upper limit of their walking speed during training may further improve walking outcomes.

These results build on prior evidence which has demonstrated training speed and/or step count (a proxy of exercise repetition in walking interventions) have a stronger association with walking capacity outcomes than training heart rate.^8,17,24^ The HIT-Stroke randomized clinical trial found training speed and training step count were significant mediators of the change in 6MWT, regardless of if participants received a high-intensity interval walking intervention or moderate-intensity continuous walking intervention.^17,24^ Similar to this current analyses, the prior analysis also found heart rate was not a significant mediator of 6MWT outcomes. As the present analysis incorporates a much larger (*n* = 129 vs. 55) sample, and accounts for various demographic variables, clinical factors, and exercise volume, it further emphasizes that training speed may be a more valuable metric of exercise intensity to improve walking capacity in people with chronic stroke. Step counts, which are correlated to walking capacity outcomes in people with chronic stroke,^8^ were not recorded during training sessions in this parent clinical trial.^9,26^ However, as training time remained constant in this protocol, an increase in walking speed in the intervention would suggest a concurrent increase in training step counts.

Previous evidence indicates walking capacity may be most strongly influenced by neuromuscular deficits in people with chronic stroke.^36,37,44^ If the neuromuscular system is the “weakest link”, walking interventions could target these deficits by focusing on challenging walking speed in training.^45–47^ Our results support this theory by suggesting a slight change in training speed, from self-selected to fastest walking speed, may lead to greater improvements in walking capacity outcomes. Furthermore, the strength of training speed as the metric of exercise intensity stands even while accounting for various other covariates known to impact walking capacity in people with chronic stroke. This is especially important as previous work which found associations between exercise intensity and walking outcomes, did not account for other variables. Therefore, the previously found association between heart rate intensity and walking capacity outcomes may have been due to other factors.

Surprisingly, training heart rate was not a significant predictor of walking capacity outcomes. The average training heart rate (64% HRR) exceeded the ACSM threshold of “vigorous intensity” exercise (> 60% HRR) and the recommended intensity of aerobic exercise for people with chronic stroke.^7,39,48,49^ There is likely a relationship between training speed and training heart rate which may partially explain why previous walking exercise interventions which aimed for moderate-to-high heart rate intensities demonstrated significant changes in walking capacity.^7^ However, recent work has questioned the use of heart rate to guide exercise intensity, noting it may not be specific enough to elicit desired outcomes.^25,50^ This is because heart rate is an indirect metric of intensity, and can easily be impacted by factors unrelated to exercise.^10^ For example, anxiety, poor sleep, or caffeine intake could influence an individual’s training heart rate. Despite these influences on heart rate being completely agnostic to the exercise intervention, they would influence this as a metric being used to quantify training intensity. It is possible that heart rate is too indirect of a measurement of intensity, particularly in people with chronic stroke who may have concurrent cardiovascular comorbidities, use beta-blocker medications, or neuromuscular deficits which impact their capacity to walk. If in the context of walking the cardiovascular system is *not* the most limiting system, then heart rate may not be the optimal target of exercise intensity. As most people with stroke cite a desire to improve their walking, determining the most specific parameters of walking exercise dose to achieve this goal is paramount.^51^

### Limitations

This walking exercise protocol was designed as a continuous walking model, so it cannot directly be compared to walking protocols which used intermittent, high-intensity (dictated by either speed or heart rate) intervals. As this protocol was directed by heart rate intensities, it is unknown if participants would be able to sustain higher training walking speeds than this study (107% of SSWS) for the same training duration. Future work is needed to decipher the strength and relationship of each exercise dose parameter to optimize walking capacity outcomes.

As the cardiopulmonary exercise stress test which determined the maximal heart rate and training heart rate zones was not repeated during the intervention, the training heart rate was not adjusted throughout. This may have failed to account for potential training effects, and limited heart rate as a metric of intensity. While heart rate calculations did not adjust for beta-blockers, participants were instructed to take beta-blocker medications both at the exercise stress test and for each training session to account for beta-blocker use. However, prior work has demonstrated individuals with chronic stroke may not reach a cardiovascular maximum during cardiopulmonary exercise stress tests,^52^ which could have limited the average heart rate intensities achieved by the sample.

Lastly, while improvements in self-selected walking speed occurred from the baseline to post-intervention timepoint, it is unknown when these improvements occurred within the intervention. Therefore, in this secondary analysis, we were unable to adjust the training speed to reflect potential changes in walking speed that were occurring throughout the intervention. Future studies could assess walking speed throughout the intervention to probe when these changes are occurring. Probing when changes in the neuromuscular and cardiovascular system are occurring within a walking intervention may elucidate which (or when) each system is the primary driver of walking capacity changes. Lastly, while these results, along with recent results from an independent cohort of people with chronic stroke, suggest training speed may be a stronger measure of exercise intensity, this may not be true for all within the chronic stroke population.^24^

## CONCLUSIONS

Walking exercise interventions, primarily conducted at moderate-to-high heart rate intensities, have demonstrated significant increases in walking capacity in people with chronic stroke, yet there remains wide variability in participant response.^6^ Results from this large cohort of people with chronic stroke who underwent a walking exercise intervention indicate training speed, but not training heart rate, significantly predicts changes in walking outcomes, after accounting for other important factors. Using training speed to guide exercise intensity, and challenging people with stroke to train at their fastest walking speed, may increase walking capacity gains in rehabilitation.

## LIST OF NON-STANDARD ABBREVIATIONS

10mWT: 10-meter walk test
6MWT: Six-Minute Walk Test
ABC: Activities-specific Balance Confidence scale
ACSM: American College of Sports Medicine
CCI: Charlson Co-morbidity Index
FAST: high-intensity treadmill training
FWS: fastest walking speed
HRR: Heart Rate Reserve
MHR: maximum heart rate
PROWALKS: Promoting Recovery Optimization of Walking Activity in Stroke
RHR: resting heart rate
SAM: step-activity behavioral intervention
SSWS: self-selected walking speed
TSS: time since stroke

**Supplemental Table 1:** Heart Rate Reserve as a Predictor of Change in 6MWT.

| Parameter | *B* | Std. Error | *t* | Sig. | 95% CI |
| --- | --- | --- | --- | --- | --- |
| Intercept | 68.639 | 47.715 | 1.439 | 0.153 | -25.850 – 163.129 |
| Sex | 5.277 | 8.951 | 0.590 | 0.557 | -12.449 – 23.003 |
| Age | -0.445 | 0.314 | -1.419 | 0.158 | -1.066 – 0.1076 |
| TSS | -0.062 | 0.053 | -1.186 | 0.238 | -0.166 – 0.042 |
| Baseline 6MWT | -0.083 | 0.041 | -2.043 | 0.043 | -0.164 – -0.003 |
| ABC | 0.297 | 0.309 | 0.961 | 0.338 | -0.314 – 0.908 |
| CCI | -1.054 | 2.655 | -0.397 | 0.692 | -6.312 – 4.203 |
| Exercise Volume | 0.036 | 0.029 | 1.234 | 0.220 | -0.022 – 0.094 |
| Heart Rate Reserve | -0.390 | 0.412 | -0.948 | 0.345 | -1.205 – 0.425 |

*Robust Errors were used. TSS = time since stroke; 6MWT = Six-Minute Walk Test, ABC = Activities Balance Confidence Scale; CCI = Charlson Co-morbidity Index; FWS = Fastest Walking Speed. Significance set at $p < 0.05$ .*

## Data Availability

All data produced in the present study are available upon reasonable request to the authors. All data from the parent randomized controlled trial can be found online at the NICHD DASH Repository.

https://dash.nichd.nih.gov/study/425019

